# Cancer burden on piecemeal endoscopic resection of early adenocarcinoma in Barrett’s oesophagus correlates with the risk of neoplastic recurrence

**DOI:** 10.1101/2025.08.29.25334287

**Authors:** Grace J Hattersley, Andreas Hadjinicolaou, Andrea Sorge, Daniel Conceicao, Sally Pan, Vijay Sujendran, Andrea Brown, Philip Kaye, Pradeep Mundre, Jacobo Ortiz-Fernández-Sordo, Massimiliano di Pietro

## Abstract

**Background and study aims:** Endoscopic resection (ER) is curative for early-stage oesophageal adenocarcinoma (OAC) without high-risk features. Piecemeal endoscopic mucosal resection (pEMR) prevents assessment of lateral margins, complicating risk estimation for neoplastic recurrence. We investigated risk factors for residual and recurrent OAC post-pEMR.

**Methods:** We performed a longitudinal study of two independent patient cohorts: the test cohort, who underwent piecemeal or en-bloc ER, (n=138) and the validation cohort, treated with pEMR only (n=89). Inclusion criteria were: OAC stage T1a or low-risk T1b, no lympho-vascular invasion, and R0 resection. The primary outcome was residual OAC at first post-ER endoscopy, and secondary outcomes were residual high-grade dysplasia (HGD), recurrence of neoplasia at any post-ER endoscopy, and remission of neoplasia and metaplasia at most recent endoscopy.

**Results:** In the test cohort, the incidence of HGD recurrence was higher in patients treated with pEMR versus en-bloc ER (*p=*0.021). The percentage of pEMR specimens with OAC was an independent risk factor for residual OAC at the first post-pEMR endoscopy (OR for a 10% increase=1.21, CI=1-1.46, *p*=0.044). A 50% cut-off of involved pEMR specimens was optimal to predict residual OAC (specificity=0.69, sensitivity=0.63). Rates of residual (*p*=0.02) and recurrent (*p*=0.0024) OAC were higher when >50% of pEMR specimens were involved by OAC. In the validation cohort, recurrent OAC was also more frequent when cancer burden was >50% (*p*=0.013).

**Conclusions:** High OAC burden on pEMR specimens correlates with the risk of residual OAC. Post-pEMR site check before endoscopic ablation is recommended if more than 50% of pEMR specimens show OAC.

## Introduction

There were an estimated 85,700 global cases of oesophageal adenocarcinoma (OAC) in 2020.^1^ This global burden has been increasing steadily since the 1970s, with a particular prevalence in male populations in the Western world, including Europe, North America and Australia.^2^ Indeed, in these areas, incidence of OAC has surpassed that of oesophageal squamous cell carcinoma.

OAC typically develops on a background of Barrett’s oesophagus (BO).^3^ Whilst unselected screening is not typically indicated, in certain high-risk groups such as patients with gastro-oesophageal reflux disease (GORD) and additional risk factors including white race, male sex and obesity, targeted screening for BO may be considered.^4–6^ For patients with diagnosed BO, endoscopic surveillance using high-definition systems with mapping biopsies according to the Seattle protocol is currently recommended by many professional guidelines, with the frequency dependent on the length of the BO segment. If visible abnormalities are detected, targeted biopsies of the most suspicious part of the lesion should be taken. Overall, the goal of endoscopic surveillance is detection of early stage, low-risk OAC, which is more amenable to endoscopic treatment.

T1a OAC with invasion confined to the muscularis mucosae is an absolute indication for endoscopic resection (ER). There is also a relative indication for ER in low-risk T1b OAC, if invasion is limited to the superficial submucosa, differentiation is well or moderate and there is no evidence of lympho-vascular invasion, because of the increasing evidence of low incidence of metastatic disease.^5,6^ At this early stage, ER is considered curative, provided the resection margins are histologically free of disease (R0), and is equally as effective as oesophagectomy whilst being associated with fewer adverse events.^7^ Ablation therapy is subsequently performed post-ER to eradicate the remaining BO epithelium and to reduce the risk of metachronous disease.^8^

Endoscopic mucosal resection (EMR) is the recommended ER technique for early-stage OAC of less than 20mm in diameter and for larger BO-associated flat dysplastic lesions.^9^ Given the typical size of an EMR specimen is 15mm, a piecemeal EMR (pEMR) is often required to ensure complete lesion resection. For early-stage OAC that is larger than 20mm in diameter or lesions with suspected superficial submucosal invasion, endoscopic submucosal dissection (ESD) is recommended. However, previous studies have shown that expert endoscopists poorly predict submucosal invasion based on endoscopic features and suboptimally agree on Paris classification.^10,11^ Limited studies have been performed comparing the efficacy of these two techniques for OAC,^12^ however it is of note that ESD is associated with longer procedural time and the absolute requirement of general anaesthetic.^13^

The goal of an ER is to remove all cancerous tissue and to clear the field of visible surrounding dysplasia. In BO, this can be difficult to fully recognise as the epithelium can bear foci of flat intramucosal cancer. Therefore, ER poses the risk of residual or recurrent disease due to dysplastic margins being overlooked, leading to inadequate resection.^7^ pEMR may have a higher risk of such recurrence as it is not possible to assess lateral margins histologically post-resection, and stratification for the risk of residual and recurrent adenocarcinoma is more challenging compared to en-bloc ER.^14,15^ Importantly, performing ablation therapy in the presence of residual OAC can jeopardise outcomes by burying disease foci and enabling deeper proliferation of dysplastic lesions.^16,17^ However, no studies to date have developed a method to inform the risk of residual OAC post-pEMR. The primary aim of this study was to identify risk factors for residual OAC at the first post-pEMR endoscopy in our patient cohort. The secondary aims were to generate a clinical template that could be used to risk stratify patients treated with pEMR for OAC.

## Materials and methods

### Study design and participants

We performed a longitudinal cohort study of patients treated for early-stage oesophageal adenocarcinoma (OAC) by endoscopic resection (ER) at Cambridge University Hospitals (CUH) between 29/03/2006 and 02/11/2023 as part of the ethically approved Cambridge registry study (LREC01/149). We included 138 patients, of which 108 patients were treated with piecemeal endoscopic mucosal resection (pEMR), 22 with en-bloc endoscopic mucosal resection (eEMR) and 8 with en-bloc endoscopic submucosal dissection (ESD). These patients are referred to as the test cohort. Inclusion criteria were: intramucosal (T1a) OAC or low-risk submucosal (T1bsm1) OAC (invasion into the submucosal layer limited to 500 microns, good to moderate differentiation); no evidence of lympho-vascular invasion (LVI-); R0 resection, defined as free vertical margins for pEMR and free vertical and radial margins for en-bloc ER; and adequate post-ER follow-up, defined as diagnostic endoscopy with biopsies or repeat ER in case of visible lesions prior to ablation therapy. This last inclusion criteria was possible as performing a site-check prior to radiofrequency ablation (RFA) is routine practice at CUH. We excluded any cases of high-grade dysplasia (HGD) only or high-risk T1b OAC (poor differentiation/grade 3 or T1bsm2-3), and any patients with inadequate information reported post-ER.

The validation independent cohort included 89 patients treated with pEMR at Nottingham University Hospitals (NUH) between 02/01/2012 and 26/07/2024 (n=57) and at Bradford Teaching Hospitals (BTH) between 01/05/2014 and 13/11/2015 (n=32). Inclusion and exclusion criteria were as above, except a pre-ablation site check with biopsies was not required for inclusion. This was because ablation was typically performed at the immediate post-pEMR endoscopy at NUH and BTH, with biopsies only taken in cases of suspicious lesions or, at BTH, if the lateral pEMR margin was positive for invasive cancer. Patients who did not have biopsies from the immediate post-pEMR endoscopy were excluded from relevant analysis if they went on to have recurrence of OAC or HGD as it would not be possible to distinguish between residual and recurrent OAC. All patients in the test and validations cohorts provided informed consent to participate in research.

### Procedures

pEMR was performed with Duette device (Cook medical) typically under conscious sedation with prior marking by snare tip. Decision to perform submucosal injection and endoscopic mucosal resection (EMR) margin coagulation was at discretion of the treating physician. From 2020 at NUH, procedures were performed under deep propofol sedation or general anaesthetic, at the discretion of the anaesthetist. As part of this study, we were unable to ascertain completion of the marked area resection. Following resection, pEMR specimens were individually pinned on cardboard. En-bloc resection was performed either with EMR Duette kit, or with ESD, which was introduced at CUH in 2017. ESD was always performed under general anaesthesia. Decision for EMR vs ESD was taken based on early cancer lesion factors with ESD preferred for sessile lesions (Paris 0-Is), lesions with depression (Paris 0-IIc) or lesion that could not be sucked in the Duette cap. At NUH, ESD was also preferred in patients with history of prior endotherapy. Given that the cohort partly preceded the most recent European Society of Gastrointestinal Endoscopy (ESGE) guidelines, the size of the lesion was typically not a determinant for treatment decision. Reporting of the ER specimen was performed according to the Vienna classification. The pathologist reported the number of specimens involved by OAC in case of pEMR.

Following a potentially curative ER, patients received a site check at 2-3 months prior to ablation therapy such as RFA. During this procedure, biopsies would be taken on the EMR scar or a subsequent ER would be performed in case of clear evidence of residual neoplasia. Indication for RFA was flat Barrett’s post-ER with no evidence of visible neoplastic lesion at post-ER endoscopic follow up. Cases without biopsies or repeat EMR before ablation therapy were excluded.

### Outcomes

The primary outcome was histological diagnosis of residual OAC in biopsies taken from the first post-ER endoscopy. Secondary outcomes were: any residual neoplasia in biopsies taken from the first post-ER endoscopy; histological evidence of recurrence of OAC and HGD identified in biopsies taken from any post-ER endoscopy (including the first post-ER endoscopy); and complete remission of OAC, dysplasia and intestinal metaplasia (IM) in biopsies taken from the most recent endoscopy. A stepwise progression model to OAC from IM to low-grade dysplasia (LGD) to HGD and finally OAC was followed, whereby any cases of residual, recurrent or failed remission of a grade of neoplasia (e.g. OAC) were also considered to be cases of lower grades of neoplasia/metaplasia (e.g. HGD, LGD and IM).

### Analysis

Descriptive statistics are expressed as medians (interquartile range) or frequency of observations with percentages. Patients in the CUH cohort who did not have alcohol consumption, ever smoker status, body mass index (BMI) or endoscopic lesion size reported, and patients in the validation cohort who either had no biopsies taken at the most recent endoscopy or where pathology at the most recent endoscopy was not available were excluded from the relevant analyses. For discrete variables, chi-squared tests were applied to contingency tables where all observations were greater than 5, otherwise Fisher’s exact test was applied. For continuous variables, Shapiro-Wilk test was applied to assess for normality, followed by the Mann-Whitney test for non-parametric distributions or the Student’s T-test for parametric distributions. Kaplan-Meier analysis with a log-rank test and a univariable Cox hazard model was performed to compare recurrence over time. The percentage of pEMR specimens with OAC was calculated as the number of pEMR specimens with histological evidence of OAC divided by the number of pEMR specimens assessed histologically. Firth’s penalised logistic regression models were generated to evaluate possible risk factors for primary and secondary outcomes. Variables included in the model were selected based on previous reports in the literature, and all variables were included in the multivariable model, except where they were based on a different cohort size (BMI and lesion size) or accounted for by an alternative variable (number of pEMR specimens with OAC and total number of pEMR specimens). Receiver operating characteristics (ROC) analysis was used to identify the optimal cut-off of pEMR specimens with OAC for residual and recurrent OAC detection. *p* values of less than 0.05 were considered statistically significant. Power analysis showed that the validation cohort would require a total sample size of 79 to detect the same difference in recurrence of OAC by pEMR cut-off that was observed in the test cohort at α of 0.05 and power of 0.8 (1-β).

All statistical analysis was performed using R for Statistical Computing v4.4.1 [R core team]. Kaplan Meier analysis was completed using the survival package and ROC analysis was completed using the pROC package. Figures were plotted using the ggplot2 and survminer packages.

## Results

### The incidence of dysplastic recurrence is higher in patients treated with piecemeal compared to en-bloc resection

A total of 420 patients with records of an oesophageal ER at Cambridge University Hospitals were assessed for inclusion in the study. After exclusion criteria were applied, we included 138 individuals who received ER for early-stage oesophageal adenocarcinoma (OAC) (Table 1, Figure 1). 108 individuals were treated with piecemeal endoscopic mucosal resection (pEMR) and 30 individuals were treated with en-bloc ER (EMR n=22; ESD n=8) (Supplementary Table 1). Baseline cohort characteristics did not differ between piecemeal and en-bloc groups, except for body mass index (BMI), which was higher in individuals treated with en-bloc ER (*p*=0.023). However, the percentage of overweight patients did not differ between ER types (*p*=1). OAC resected by pEMR was more commonly of poor differentiation (*p*=0.0076).

**Figure 1:**
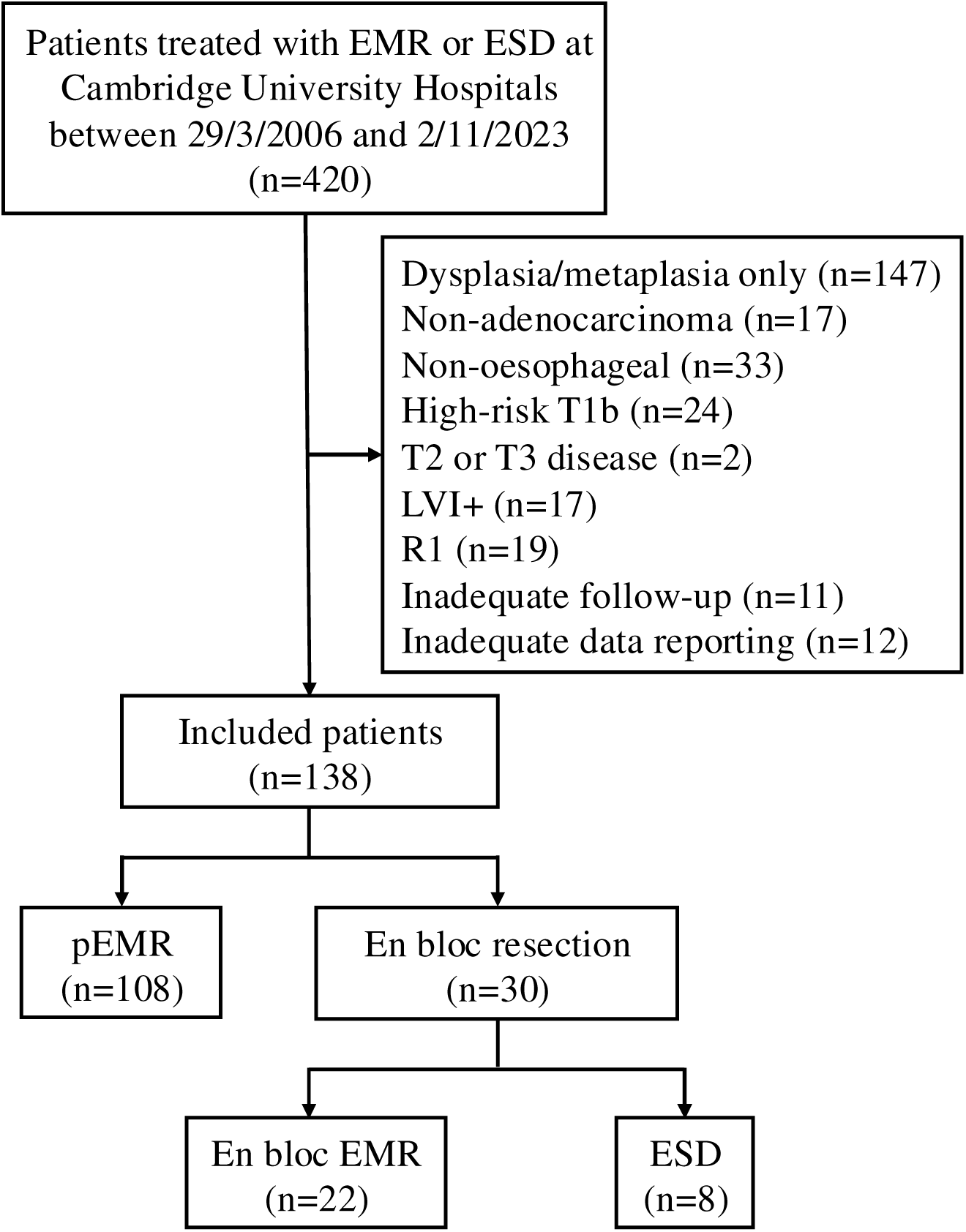
Flow chart demonstrating included and excluded patients

**Table 1:** Cohort characteristics by type of resection, pEMR or en-bloc ER (en-bloc EMR and ESD). Data are n (%) or median (IQR). Bold indicates *p*<0.05. *Based on a total cohort size of 129. **Based on a total cohort size of 110. ***Based on a total cohort size of 103. ****Based on a total cohort size of 89.

|  | Total cohort (n=138) | pEMR (n=108) | En-bloc ER (n=30) | P-value |
| --- | --- | --- | --- | --- |
| Age | 68 (64-75.75) | 69 (64-77) | 68 (64-73) | 0.3 |
| Sex (Male) | 112 (81%) | 89 (82%) | 23 (77%) | 0.65 |
| BMI* | 29 (26.3-31.3) | 28.6 (25.9-30.7) | 30.9 (27.5-35.9) | **0.023** |
| Overweight (BMI > 25)* | 110 (85%) | 86 (85%) | 24 (86%) | 1 |
| Time to first follow-up (months) | 2 (1-2) | 2 (1-2) | 2 (2-3.75) | 0.089 |
| Duration of follow-up (months) | 44 (20-70.75) | 43.5 (19.75-70.25) | 49.5 (20.25-73.25) | 0.79 |
| Ever smoker** | 76 (69%) | 56 (67%) | 20 (74%) | 0.69 |
| Alcohol excess (>14 units per week)*** | 15 (15%) | 13 (16%) | 2 (8%) | 0.51 |
| Lesion size (mm)**** | 20 (10-25) | 20 (10-25) | 10 (9-20) | 0.072 |
| Stage (T1bsm1) | 7 (5%) | 5 (5%) | 2 (7%) | 0.65 |
| Differentiation (Poor) | 7 (5%) | 6 (6%) | 1 (3%) | **0.0076** |
| Histology at first follow-up |  |  |  |  |
| OAC | 25 (18%) | 19 (18%) | 6 (20%) | 0.97 |
| HGD | 45 (33%) | 39 (36%) | 6 (20%) | 0.15 |
| LGD | 59 (43%) | 49 (45%) | 10 (33%) | 0.33 |
| IM | 118 (86%) | 95 (88%) | 23 (77%) | 0.21 |
| Recurrence within one year |  |  |  |  |
| OAC | 35 (25%) | 28 (26%) | 7 (23%) | 0.96 |
| HGD | 55 (40%) | 48 (44%) | 7 (23%) | 0.06 |
| Recurrence anytime |  |  |  |  |
| OAC | 36 (26%) | 29 (27%) | 7 (23%) | 0.88 |
| HGD | 60 (43%) | 53 (49%) | 7 (23%) | **0.021** |
| Histology at last follow-up |  |  |  |  |
| OAC | 11 (8%) | 10 (9%) | 1 (3%) | 0.46 |
| HGD | 14 (10%) | 13 (12%) | 1 (3%) | 0.3 |
| LGD | 23 (17%) | 20 (19%) | 3 (10%) | 0.41 |
| IM | 61 (44%) | 49 (45%) | 12 (40%) | 0.75 |

We assessed the incidence of residual OAC and HGD at the first post-ER endoscopy after a median interval of 2 months (Interquartile range (IQR) 1-2), and the development of OAC and HGD recurrence over a median follow-up time of 44 months (IQR 20-70.75). Altogether, 36 patients exhibited OAC on biopsies taken at any post-ER, of which 25 (69%) occurred at the immediate post-ER endoscopy. Moreover, HGD was identified in biopsies taken at any post-ER endoscopy in 60 individuals, of which 45 (75%) were diagnosed at the first post-ER endoscopy. The rate of recurrence of HGD over the total follow-up period was higher in individuals treated with pEMR compared to en-bloc ER (*p*=0.021, Table 1). Kaplan Meier analysis confirmed that the cumulative incidence of HGD recurrence over a three-year period was higher in patients treated with pEMR (*p*=0.028, HR 2.33, 95% CI 1.06-5.14; Figure 2). We hypothesised that this could be due to dysplastic margins being overlooked and thus left in situ because of the lack of circumferential incision in pEMR. Since pEMR prevents assessment of lateral resection margins, we investigated alternative factors that could be used to identify patients at risk of residual OAC post-pEMR.

**Figure 2:**
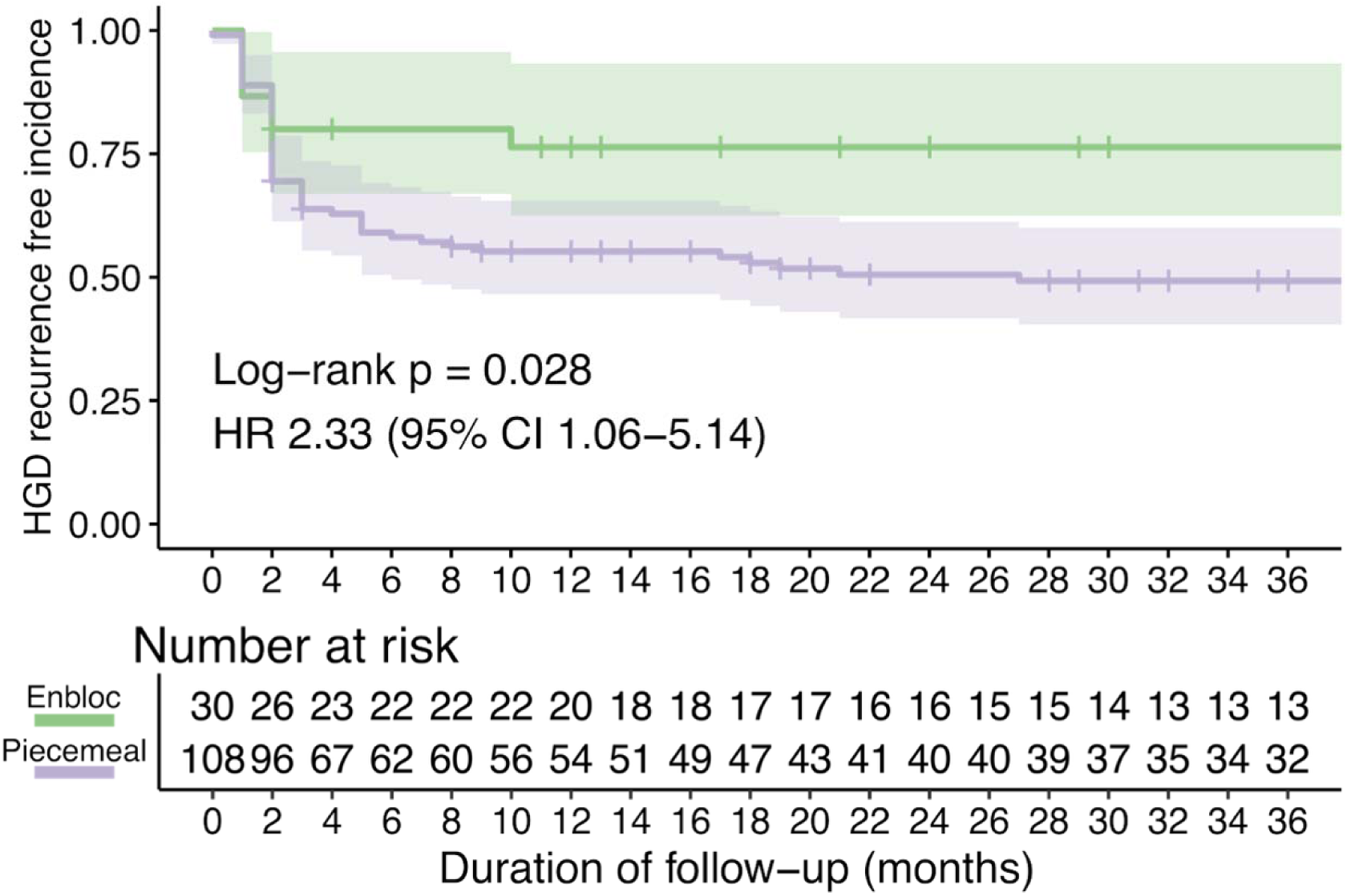
OAC recurrence-free survival of patients post-ER by resection type over a three-year period. Time measured from date of first endoscopic resection. Shaded regions represent 95% confidence intervals; crosses denote censored patients. The hazard ratio (HR) and *p* value calculated using a log rank test represent the comparison between individuals treated with pEMR and en-bloc ER.

### The percentage of pEMR specimens with OAC is an independent risk factor for residual OAC at the first post-pEMR endoscopy

We generated a multivariable logistic regression model to identify clinical factors that influenced the risk of residual OAC post-pEMR (Table 2). This revealed that the percentage of pEMR specimens with OAC at the initial resection was an independent risk factor for residual OAC at the first post-pEMR endoscopy (*p*=0.045). On univariable analysis, we identified that this was likely due to an increase in the absolute number of pEMR specimens with OAC (*p*=0.042) rather than due to a reduction in the number of pEMR specimens taken (*p*=0.27). Similarly, the percentage of pEMR specimens with OAC at the initial resection was an independent risk factor for recurrence of OAC identified at any post-pEMR endoscopy (*p*=0.018; Supplementary Table 2). However, the percentage of pEMR specimens with OAC was not a risk factor for residual dysplasia at the first post-pEMR endoscopy (HGD: *p*=0.37; LGD: *p*=0.78), or recurrence of HGD identified at any post-pEMR endoscopic follow-up (*p*=0.23). Similarly, remission of OAC (*p*=0.27), OAC and HGD (*p*=0.75) and any neoplasia (OAC, HGD and LGD; *p*=0.84) at the most recent endoscopy were not associated with the percentage of pEMR specimens with OAC. Of note, we did not identify any correlation between the percentage of pEMR specimens with OAC and the absolute number of piecemeal resections (R^2^ = 0.011, *p*=0.015), further supporting this variable as an independent risk factor for residual and recurrent OAC.

**Table 2:**
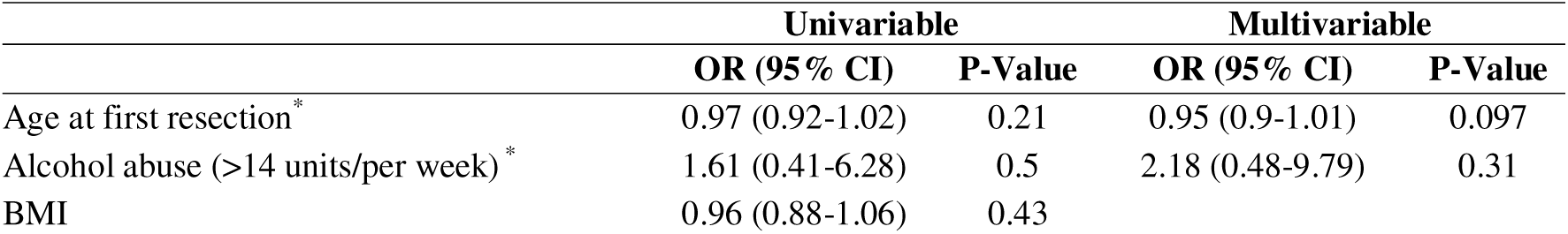
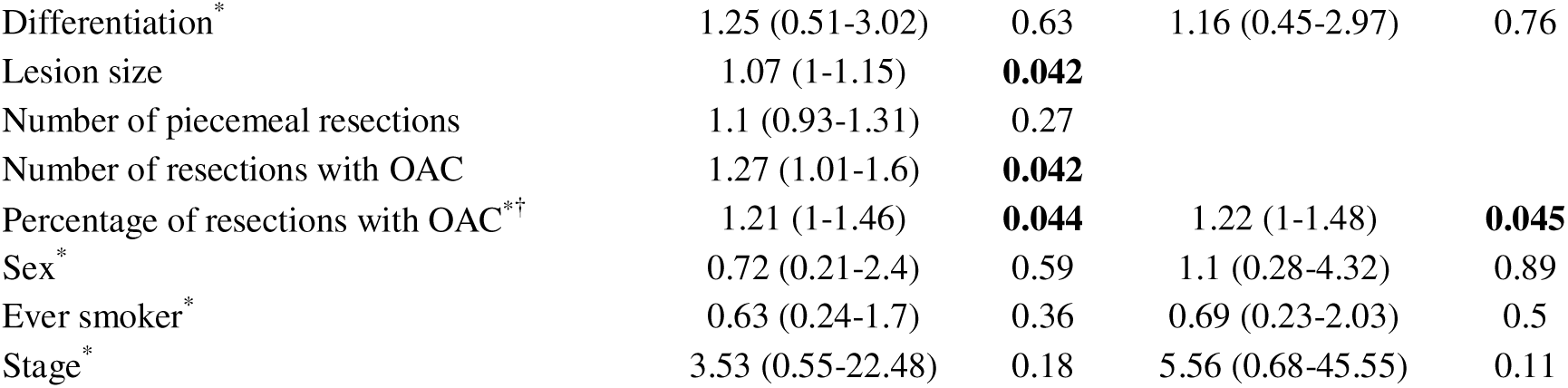
Univariable and multivariable logistic regression model for residual OAC at first post-pEMR endoscopy. *Inclusion in the multivariable model. ^†^Odds ratio for a 10% increase in the number of pEMR specimens with cancer on histological investigation.

Given the association between the percentage of samples with OAC and residual and recurrent cancer risk, we generated receiver operating characteristics (ROC) curves to identify a threshold of pEMR specimens with OAC above which patients were more likely to experience residual and recurrent cancer. This revealed that a 53.5% cut-off of pEMR specimens with OAC at the initial resection maximised sensitivity and specificity for residual (Supplementary Figure 1A) and recurrent (Supplementary Figure 1B) OAC detection. Clinically, we determined that this would correlate to a >50% cut-off, given that a minimum of 23 specimens would be necessary to distinguish a 50% cut-off from a 53.5% cut-off.

### Patients with OAC burden of more than 50% show increased rates of residual and recurrent OAC post-pEMR

To further investigate whether this cut-off of pEMR specimens with OAC could be used to risk stratify patients, we subdivided the pEMR cohort (n=108) into patients with more than 50% cancer burden (n=40) and those with less than 50% cancer burden (n=68) (Table 3). Baseline characteristics did not differ between these two groups. We found that patients with more than 50% specimens involved by OAC at initial pEMR showed an increased rate of residual OAC at first post-pEMR endoscopy (*p*=0.02), recurrence of OAC (*p*=0.0053) and HGD (*p*=0.022) at one year, and recurrence of OAC (*p*=0.0024) and HGD (*p*=0.0062) at any post-pEMR endoscopy. This was corroborated by Kaplan Meier analysis, which identified an increased incidence of OAC recurrence in patients with OAC burden >50% over a three-year period (Figure 3, *p*=0.002). The rate of remission of OAC was also reduced in patients with a greater than 50% cancer burden (*p*=0.037), however, the rates of remission of OAC and HGD (*p*=0.1) and any neoplasia (OAC, HGD and LGD: *p*=0.28) were not affected by this cut-off.

**Figure 3:**
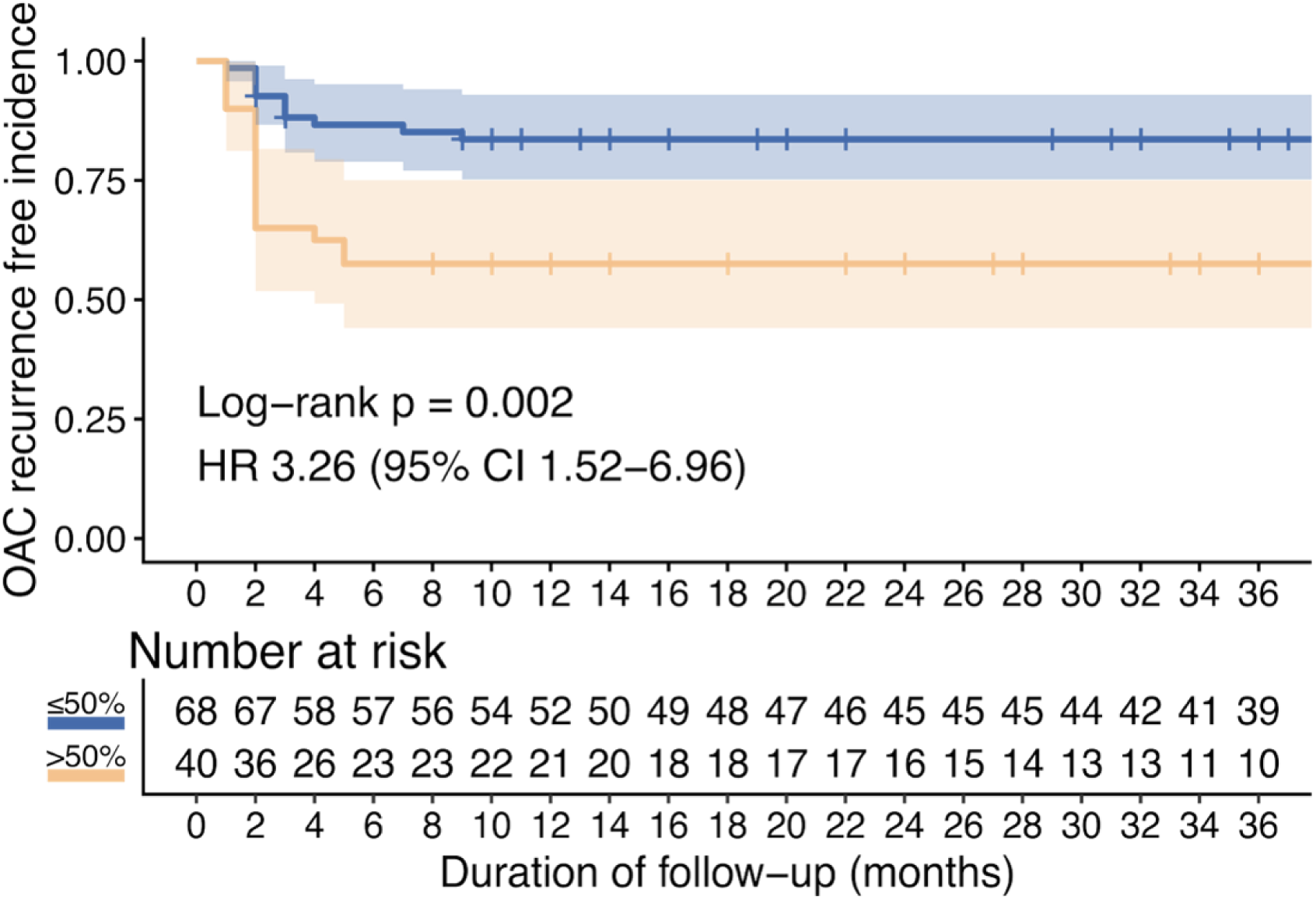
OAC recurrence-free survival of patients post-pEMR by percentage of pEMR specimens affected by OAC. Time measured from date of first endoscopic resection. Shaded regions represent 95% confidence intervals; crosses denote censored patients. The hazard ratio (HR) and p value calculated using a log rank test represents the comparison between individuals with > 50% and ≤50% of pEMR specimens with OAC.

**Table 3:**
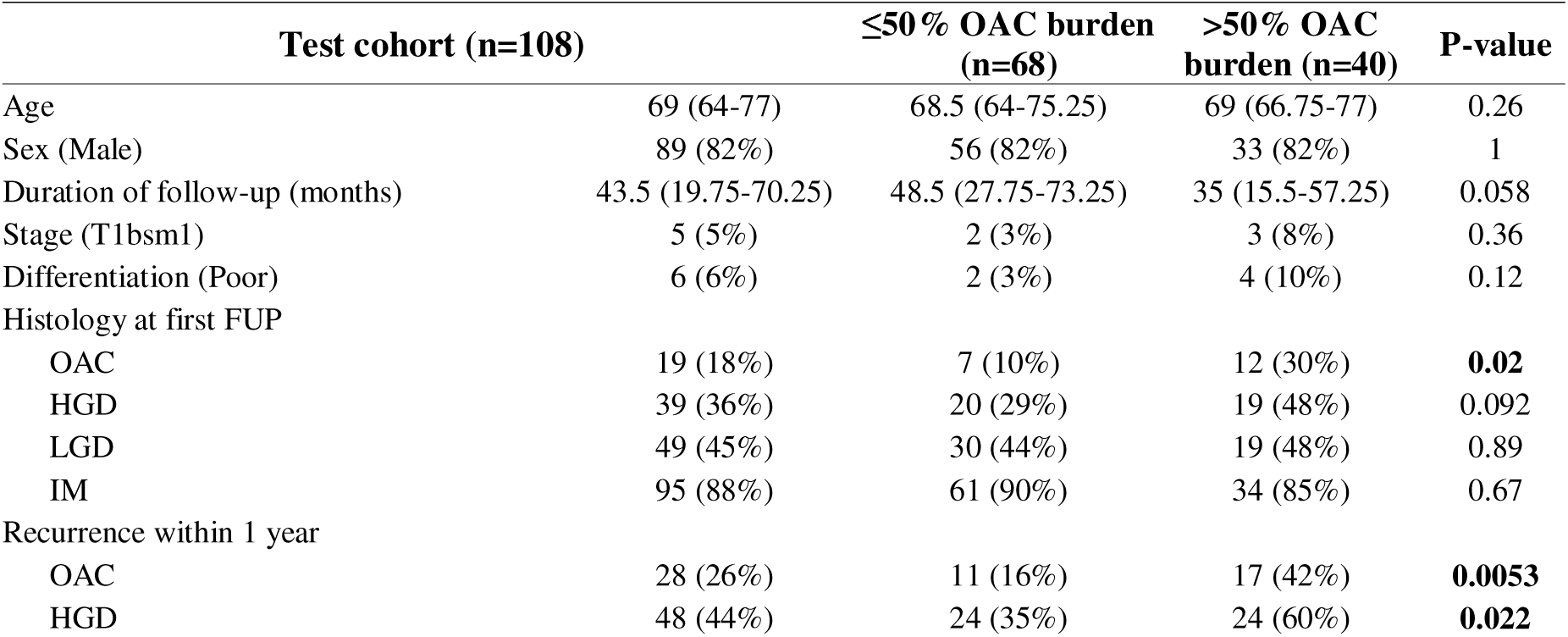
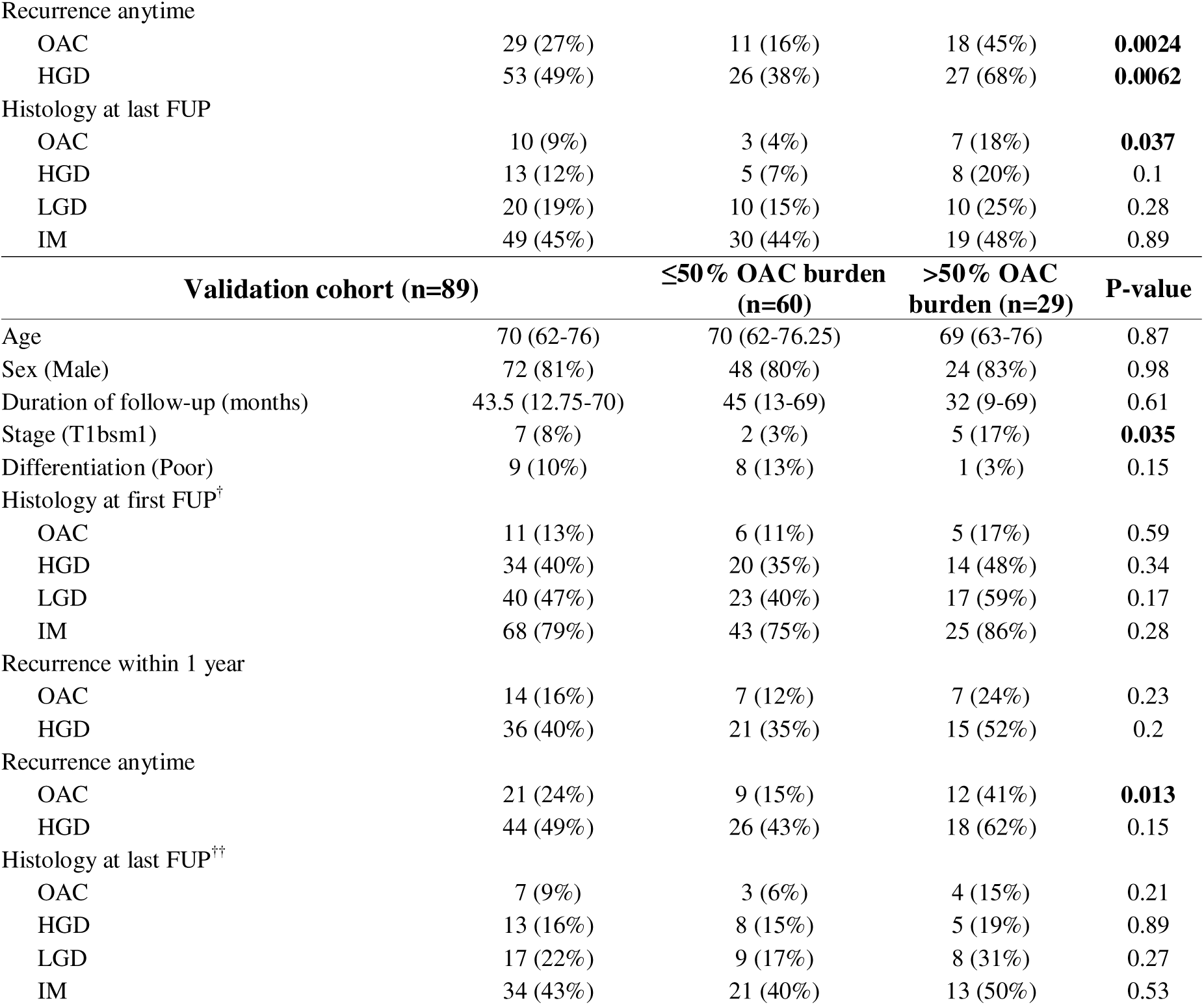
Cohort characteristics by percentage cut-off. Top panel, test cohort, derived from patients treated at a single centre; bottom panel, validation cohort, derived from patients treated at two independent centres. Data are n (%) or median (IQR). Bold indicates *p*<0.05. ^†^Based on a total cohort size of 86. ^††^Based on a total cohort size of 79.

### Validation of cancer burden cut off in an independent cohort

We further investigated the 50% cut-off of pEMR specimens with OAC in an independent patient cohort from two institutions (Table 3). Of the 89 patients in the validation cohort, 29 patients had pEMR OAC burden >50%, and 60 patients had pEMR OAC burden ≤50%. Baseline cohort characteristics were similar, except patients with a higher OAC burden were more commonly of T1bsm1 stage (*p*=0.035). We identified that there was an increased frequency of OAC recurrence in patients where more than 50% of pEMR specimens had OAC (*p*=0.013).

## Discussion

This study is, to the best of our knowledge, the first to investigate risk factors for residual OAC after pEMR. We showed that there was an increased rate of HGD recurrence post-pEMR compared to en-bloc resection modalities, including en-bloc EMR and ESD. The percentage of cancerous samples was an independent risk factor for residual-OAC post-pEMR, with a 50% cut-off of pEMR specimens with OAC representing the best threshold to identify patients at the highest risk of residual and recurrent cancer.

The results of our study have important implications in determining the optimal timing of ablation therapy following ER. Current clinical guidelines recommend ablation of the remaining Barrett’s epithelium after treatment of dysplasia or OAC with endoscopic resection. The most common ablation modality is radiofrequency ablation (RFA), which uses high temperature to a depth of 500 microns to destroy the oesophageal epithelium. This is an approved treatment modality for dysplastic BO, whereas RFA is contraindicated in the presence of intramucosal adenocarcinoma due to the risk of invasion beyond 500 microns. In this setting, RFA would cause burial of neoplastic columnar epithelium and would enable deep proliferation that evades surveillance.^16–18^ However, limited research has been performed to date to identify the optimal timing of RFA after endoscopic resection, or to assess if there is benefit in delaying RFA to prevent such subsquamous metaplasia. The EUROII study, which investigated the efficacy of RFA after EMR for T1a OAC, delayed ablation therapy until at least 2 post-EMR site checks with mapping biopsies which showed no evidence of cancer were performed. This study showed a very high rate of resolution of neoplasia and BO of 92% and 87%, respectively.^8^ Our findings suggest that patients treated with pEMR, and in particular those who have a greater than 50% cancer burden, are at higher risk for inadequate resection and recurrent dysplasia and OAC. We therefore recommend that such high-risk patients should not receive RFA at the immediate post-pEMR endoscopy to reduce the risk of burying residual OAC. Caution should generally be applied to patients treated for OAC with pEMR before starting RFA, and any visible flat lesions should be biopsied or resected.

Percentage cancer burden on pEMR represents the first approach to risk stratify patients for residual OAC and can be used to complement other models that inform risk after successful endoscopic treatment. A study from the Netherlands developed a model to predict visible dysplastic recurrence following complete eradication of Barrett’s oesophagus with OAC or dysplasia.^19^ The risk factors included in the model are new visible lesions during treatment, higher number of ER treatments, male sex, increasing Barrett’s oesophagus length, HGD or cancer at baseline, and younger patient age. Whilst this model enables a long-term follow-up plan to be tailored to an individual patient’s risk factors, the use of percentage cancer burden as a risk factor has the potential to prevent adverse patient outcomes by delaying ablation therapy.

Increased lesion size is already known to be associated with increased risk of recurrence after ER, and increasing pEMR cancer burden could be considered a surrogate of this.^20^ However, using the percentage of pEMR specimens with OAC as a risk stratification tool has several advantages. First, sizing lesions in BO can be challenging due to the field of flat dysplasia. Second, the percentage cancer burden can also reflect the endoscopists radicality in the resection. A high percentage of pEMR specimens with OAC could either reflect a limited endoscopic resection of a smaller lesion, or a neoplastic lesion of large size. In both scenarios, the theoretical risk of residual neoplasia is higher, and thus using percentage cancer burden as a risk stratification tool encompasses additional risk factors not accounted for by lesion size.

A higher rate of dysplastic recurrence has previously been reported in patients treated for OAC with EMR compared to ESD, in which the en-bloc rate was lower in patients treated with EMR, and in patients treated with a piecemeal ER compared to en-bloc ER.^20–22^ We argue that this higher rate of recurrence could be attributed to the difficulty in ensuring adequate resection of the lesion margin. Other studies comparing ESD with EMR have found no difference in the recurrence rate.^12,23,24^ Our results do not advocate against pEMR as, in our cohort, careful site check and repeat EMR if required achieved remission of OAC in more than 90% of cases. There is an ongoing randomised clinical trial comparing the efficacy of ESD and EMR for treatment of BO-related neoplasia with a primary outcome related to recurrent or residual neoplasia in the two groups at 12 months (NCT05276791). The results of this trial will be useful to validate our findings. However, this RCT will recruit patients with visible lesions, including non-invasive neoplastic lesions, which was outside of the remit of our study.

Our study has several limitations. First, whilst patients in the Cambridge cohort were recruited prospectively, retrieval of lesion size information required retrospective review of electronic healthcare records. We were unable to corroborate endoscopy results with endoscopic images, and as such lesion size information is missing for some patients and we could not retrieve sufficient data to include the Paris classification in the multivariable model. Our clinical risk factor was developed based on data from a single centre, and the resulting low cohort size may have resulted in overfitting the logistic regression model. However, the impact of this was mitigated through validation of the strongest predictor in a multicentre cohort. Power calculation to determine the minimum validation cohort size to detect the same difference in recurrence of OAC assumed the ratio of patients with more than 50% of pEMR specimens affected by OAC was the same as in the test cohort. However, data analysis revealed the validation cohort had a greater proportion of patients with less than 50% cancer burden. Hence, post-hoc power calculations revealed a power of 0.76, despite the minimum sample size of 79 patients being reached.

In summary, our study provides a novel, simple and clinically applicable parameter for risk stratification of patients with low-risk early OAC treated with pEMR and advocate for careful assessment of these patients post-ER prior to starting ablation treatment.

## Data Availability

All data produced in the present study are available upon reasonable request to the authors

## Supplementals

**Supplementary Table 1:** Cohort characteristics by type of en-bloc resection, en-bloc EMR or ESD. Data are n (%) or median (IQR). Bold indicates *p*<0.05. *Based on a total cohort size of 28. **Based on a total cohort size of 27. ***Based on a total cohort size of 24. ****Based on a total cohort size of 19.

|  | En-bloc cohort (n=30) | eEMR (n=22) | ESD (n=8) | P-value |
| --- | --- | --- | --- | --- |
| Age | 68 (64-73) | 68 (64-72.75) | 67.5 (63-75) | 0.87 |
| Sex (Male) | 23 (77%) | 16 (73%) | 7 (88%) | 0.64 |
| BMI* | 30.9 (27.5-35.9) | 29.75 (26.15-32.65) | 35.225 (30.55-39) | **0.021** |
| Overweight (BMI > 25)* | 24 (86%) | 16 (80%) | 8 (100%) | 0.29 |
| Time to first follow-up (months) | 2 (2-3.75) | 2 (1.25-2.75) | 4 (2-6) | 0.079 |
| Duration of follow-up (months) | 49.5 (20.25-73.25) | 55.5 (34-96.5) | 19 (12.5-29.25) | **0.012** |
| Ever smoker** | 20 (74%) | 16 (80%) | 4 (57%) | 0.33 |
| Alcohol excess (>14 units per week)*** | 2 (8%) | 2 (11%) | 0 (0%) | 1 |
| Lesion size (mm)**** | 10 (9-20) | 10 (7-10) | 25 (15-41.25) | **0.00083** |
| Stage (T1bsm1) | 2 (7%) | 1 (5%) | 1 (12%) | 0.47 |
| Differentiation (Poor) | 1 (3%) | 1 (5%) | 0 (0%) | 1 |
| Histology at first follow-up |  |  |  |  |
| OAC | 6 (20%) | 6 (27%) | 0 (0%) | 0.16 |
| HGD | 6 (20%) | 6 (27%) | 0 (0%) | 0.16 |
| LGD | 10 (33%) | 10 (45%) | 0 (0%) | **0.029** |
| IM | 23 (77%) | 20 (91%) | 3 (38%) | **0.0067** |
| Recurrence within one year |  |  |  |  |
| OAC | 7 (23%) | 7 (32%) | 0 (0%) | 0.14 |
| HGD | 7 (23%) | 7 (32%) | 0 (0%) | 0.14 |
| Recurrence anytime |  |  |  |  |
| OAC | 7 (23%) | 7 (32%) | 0 (0%) | 0.14 |
| HGD | 7 (23%) | 7 (32%) | 0 (0%) | 0.14 |
| Histology at last follow-up |  |  |  |  |
| OAC | 1 (3%) | 1 (5%) | 0 (0%) | 1 |
| HGD | 1 (3%) | 1 (5%) | 0 (0%) | 1 |
| LGD | 3 (10%) | 3 (14%) | 0 (0%) | 0.54 |
| IM | 12 (40%) | 9 (41%) | 3 (38%) | 1 |

**Supplementary Table 2:**
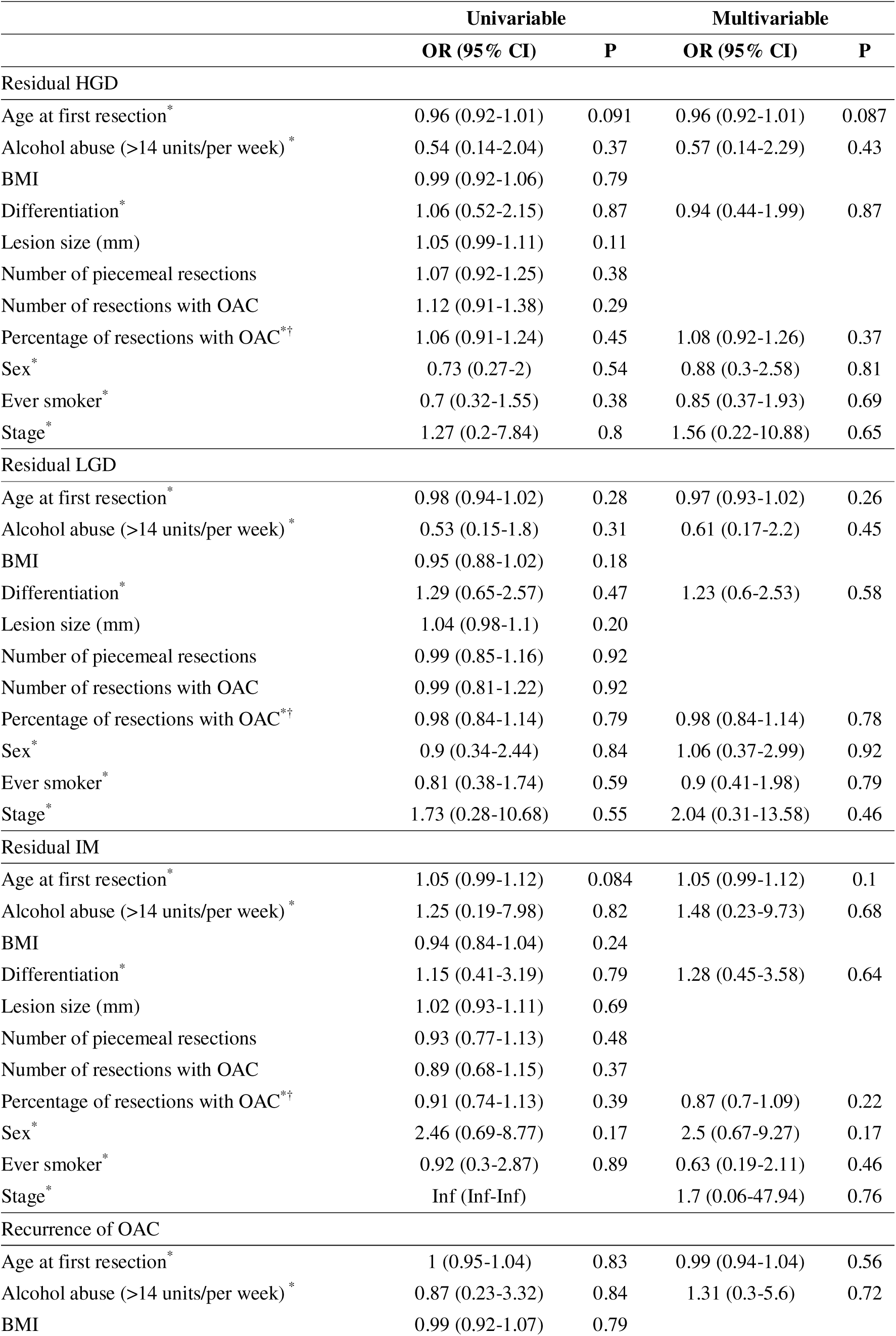
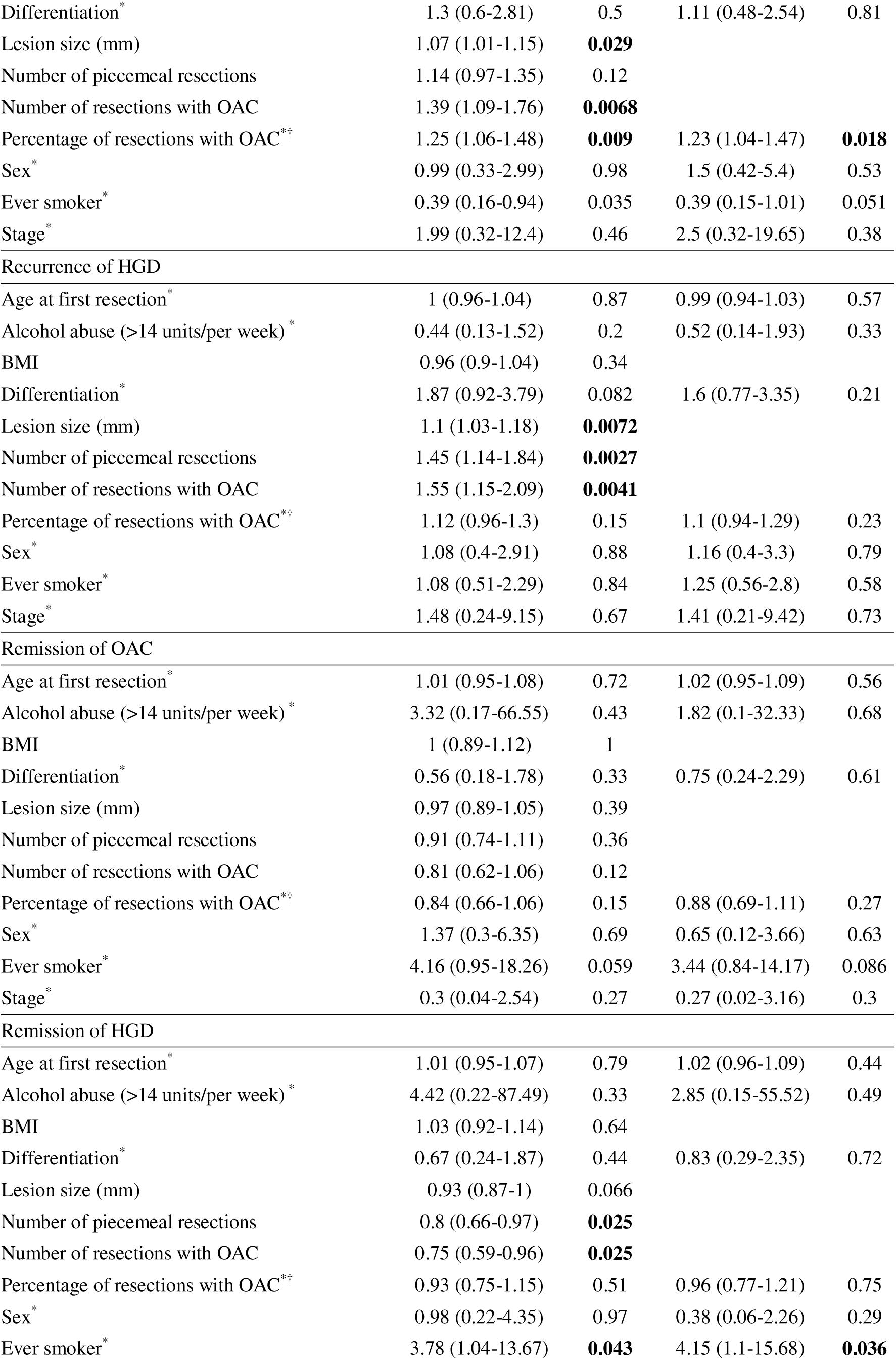
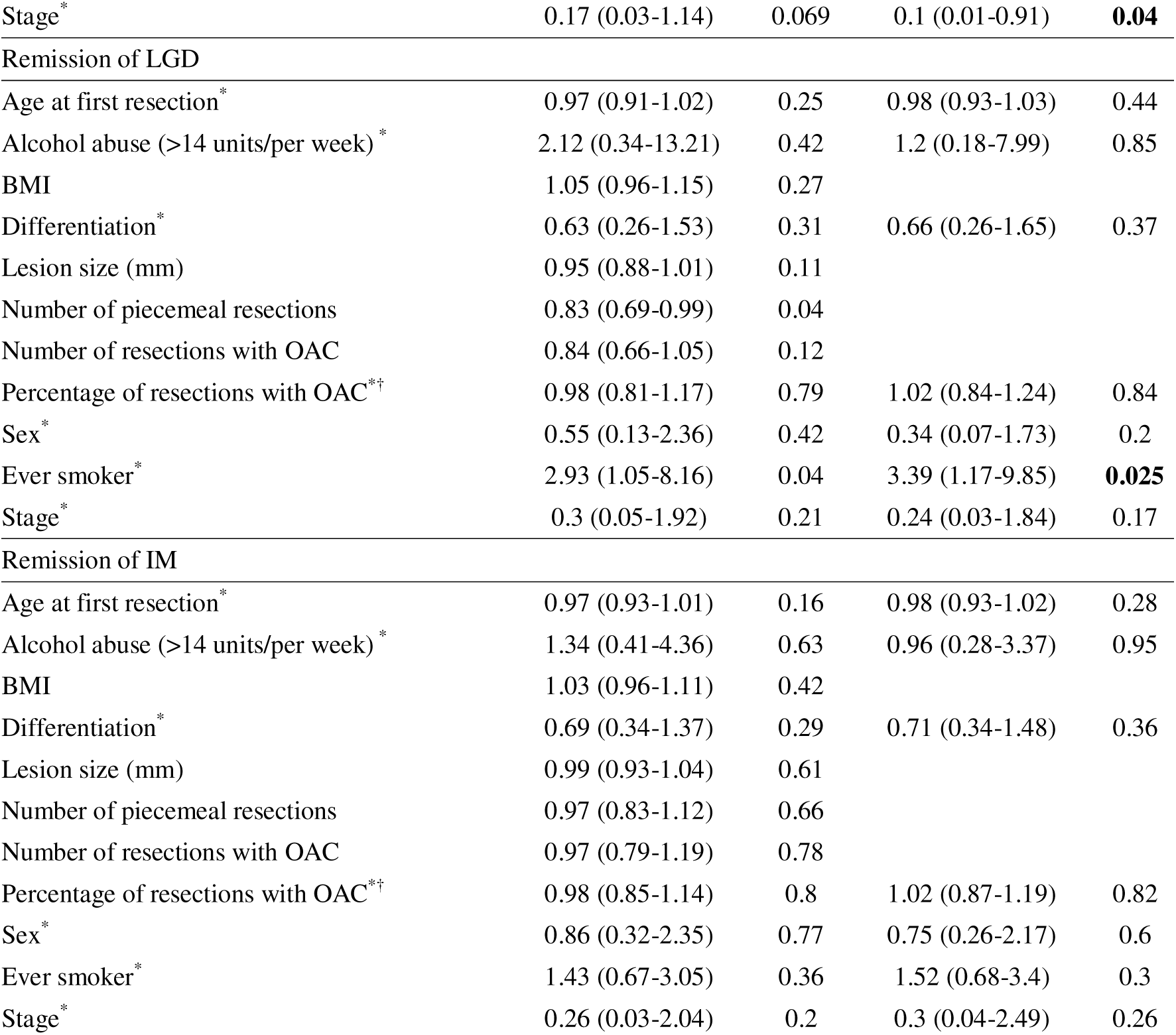
Logistic regression model for residual HGD at the first post-pEMR endoscopy, recurrence of OAC and identified at any future endoscopy and remission of OAC, HGD and IM identified at most recent endoscopy. *Inclusion in the multivariable model. ^†^Odds ratio for a 10% increase in the number of pEMR specimens with cancer on histological investigation.

|  | Univariable |  | Multivariable |  |
| --- | --- | --- | --- | --- |
|  | OR (95% CI) | P | OR (95% CI) | P |
| Residual HGD |  |  |  |  |
| Age at first resection* | 0.96 (0.92-1.01) | 0.091 | 0.96 (0.92-1.01) | 0.087 |
| Alcohol abuse (>14 units/per week) * | 0.54 (0.14-2.04) | 0.37 | 0.57 (0.14-2.29) | 0.43 |
| BMI | 0.99 (0.92-1.06) | 0.79 |  |  |
| Differentiation* | 1.06 (0.52-2.15) | 0.87 | 0.94 (0.44-1.99) | 0.87 |
| Lesion size (mm) | 1.05 (0.99-1.11) | 0.11 |  |  |
| Number of piecemeal resections | 1.07 (0.92-1.25) | 0.38 |  |  |
| Number of resections with OAC | 1.12 (0.91-1.38) | 0.29 |  |  |
| Percentage of resections with OAC*† | 1.06 (0.91-1.24) | 0.45 | 1.08 (0.92-1.26) | 0.37 |
| Sex* | 0.73 (0.27-2) | 0.54 | 0.88 (0.3-2.58) | 0.81 |
| Ever smoker* | 0.7 (0.32-1.55) | 0.38 | 0.85 (0.37-1.93) | 0.69 |
| Stage* | 1.27 (0.2-7.84) | 0.8 | 1.56 (0.22-10.88) | 0.65 |
| Residual LGD |  |  |  |  |
| Age at first resection* | 0.98 (0.94-1.02) | 0.28 | 0.97 (0.93-1.02) | 0.26 |
| Alcohol abuse (>14 units/per week) * | 0.53 (0.15-1.8) | 0.31 | 0.61 (0.17-2.2) | 0.45 |
| BMI | 0.95 (0.88-1.02) | 0.18 |  |  |
| Differentiation* | 1.29 (0.65-2.57) | 0.47 | 1.23 (0.6-2.53) | 0.58 |
| Lesion size (mm) | 1.04 (0.98-1.1) | 0.20 |  |  |
| Number of piecemeal resections | 0.99 (0.85-1.16) | 0.92 |  |  |
| Number of resections with OAC | 0.99 (0.81-1.22) | 0.92 |  |  |
| Percentage of resections with OAC*† | 0.98 (0.84-1.14) | 0.79 | 0.98 (0.84-1.14) | 0.78 |
| Sex* | 0.9 (0.34-2.44) | 0.84 | 1.06 (0.37-2.99) | 0.92 |
| Ever smoker* | 0.81 (0.38-1.74) | 0.59 | 0.9 (0.41-1.98) | 0.79 |
| Stage* | 1.73 (0.28-10.68) | 0.55 | 2.04 (0.31-13.58) | 0.46 |
| Residual IM |  |  |  |  |
| Age at first resection* | 1.05 (0.99-1.12) | 0.084 | 1.05 (0.99-1.12) | 0.1 |
| Alcohol abuse (>14 units/per week) * | 1.25 (0.19-7.98) | 0.82 | 1.48 (0.23-9.73) | 0.68 |
| BMI | 0.94 (0.84-1.04) | 0.24 |  |  |
| Differentiation* | 1.15 (0.41-3.19) | 0.79 | 1.28 (0.45-3.58) | 0.64 |
| Lesion size (mm) | 1.02 (0.93-1.11) | 0.69 |  |  |
| Number of piecemeal resections | 0.93 (0.77-1.13) | 0.48 |  |  |
| Number of resections with OAC | 0.89 (0.68-1.15) | 0.37 |  |  |
| Percentage of resections with OAC*† | 0.91 (0.74-1.13) | 0.39 | 0.87 (0.7-1.09) | 0.22 |
| Sex* | 2.46 (0.69-8.77) | 0.17 | 2.5 (0.67-9.27) | 0.17 |
| Ever smoker* | 0.92 (0.3-2.87) | 0.89 | 0.63 (0.19-2.11) | 0.46 |
| Stage* | Inf (Inf-Inf) |  | 1.7 (0.06-47.94) | 0.76 |
| Recurrence of OAC |  |  |  |  |
| Age at first resection* | 1 (0.95-1.04) | 0.83 | 0.99 (0.94-1.04) | 0.56 |
| Alcohol abuse (>14 units/per week) * | 0.87 (0.23-3.32) | 0.84 | 1.31 (0.3-5.6) | 0.72 |
| BMI | 0.99 (0.92-1.07) | 0.79 |  |  |
| Differentiation* | 1.3 (0.6-2.81) | 0.5 | 1.11 (0.48-2.54) | 0.81 |
| Lesion size (mm) | 1.07 (1.01-1.15) | **0.029** |  |  |
| Number of piecemeal resections | 1.14 (0.97-1.35) | 0.12 |  |  |
| Number of resections with OAC | 1.39 (1.09-1.76) | **0.0068** |  |  |
| Percentage of resections with OAC*† | 1.25 (1.06-1.48) | **0.009** | 1.23 (1.04-1.47) | **0.018** |
| Sex* | 0.99 (0.33-2.99) | 0.98 | 1.5 (0.42-5.4) | 0.53 |
| Ever smoker* | 0.39 (0.16-0.94) | 0.035 | 0.39 (0.15-1.01) | 0.051 |
| Stage* | 1.99 (0.32-12.4) | 0.46 | 2.5 (0.32-19.65) | 0.38 |
| Recurrence of HGD |  |  |  |  |
| Age at first resection* | 1 (0.96-1.04) | 0.87 | 0.99 (0.94-1.03) | 0.57 |
| Alcohol abuse (>14 units/per week)* | 0.44 (0.13-1.52) | 0.2 | 0.52 (0.14-1.93) | 0.33 |
| BMI | 0.96 (0.9-1.04) | 0.34 |  |  |
| Differentiation* | 1.87 (0.92-3.79) | 0.082 | 1.6 (0.77-3.35) | 0.21 |
| Lesion size (mm) | 1.1 (1.03-1.18) | **0.0072** |  |  |
| Number of piecemeal resections | 1.45 (1.14-1.84) | **0.0027** |  |  |
| Number of resections with OAC | 1.55 (1.15-2.09) | **0.0041** |  |  |
| Percentage of resections with OAC*† | 1.12 (0.96-1.3) | 0.15 | 1.1 (0.94-1.29) | 0.23 |
| Sex* | 1.08 (0.4-2.91) | 0.88 | 1.16 (0.4-3.3) | 0.79 |
| Ever smoker* | 1.08 (0.51-2.29) | 0.84 | 1.25 (0.56-2.8) | 0.58 |
| Stage* | 1.48 (0.24-9.15) | 0.67 | 1.41 (0.21-9.42) | 0.73 |
| Remission of OAC |  |  |  |  |
| Age at first resection* | 1.01 (0.95-1.08) | 0.72 | 1.02 (0.95-1.09) | 0.56 |
| Alcohol abuse (>14 units/per week)* | 3.32 (0.17-66.55) | 0.43 | 1.82 (0.1-32.33) | 0.68 |
| BMI | 1 (0.89-1.12) | 1 |  |  |
| Differentiation* | 0.56 (0.18-1.78) | 0.33 | 0.75 (0.24-2.29) | 0.61 |
| Lesion size (mm) | 0.97 (0.89-1.05) | 0.39 |  |  |
| Number of piecemeal resections | 0.91 (0.74-1.11) | 0.36 |  |  |
| Number of resections with OAC | 0.81 (0.62-1.06) | 0.12 |  |  |
| Percentage of resections with OAC*† | 0.84 (0.66-1.06) | 0.15 | 0.88 (0.69-1.11) | 0.27 |
| Sex* | 1.37 (0.3-6.35) | 0.69 | 0.65 (0.12-3.66) | 0.63 |
| Ever smoker* | 4.16 (0.95-18.26) | 0.059 | 3.44 (0.84-14.17) | 0.086 |
| Stage* | 0.3 (0.04-2.54) | 0.27 | 0.27 (0.02-3.16) | 0.3 |
| Remission of HGD |  |  |  |  |
| Age at first resection* | 1.01 (0.95-1.07) | 0.79 | 1.02 (0.96-1.09) | 0.44 |
| Alcohol abuse (>14 units/per week)* | 4.42 (0.22-87.49) | 0.33 | 2.85 (0.15-55.52) | 0.49 |
| BMI | 1.03 (0.92-1.14) | 0.64 |  |  |
| Differentiation* | 0.67 (0.24-1.87) | 0.44 | 0.83 (0.29-2.35) | 0.72 |
| Lesion size (mm) | 0.93 (0.87-1) | 0.066 |  |  |
| Number of piecemeal resections | 0.8 (0.66-0.97) | **0.025** |  |  |
| Number of resections with OAC | 0.75 (0.59-0.96) | **0.025** |  |  |
| Percentage of resections with OAC*† | 0.93 (0.75-1.15) | 0.51 | 0.96 (0.77-1.21) | 0.75 |
| Sex* | 0.98 (0.22-4.35) | 0.97 | 0.38 (0.06-2.26) | 0.29 |
| Ever smoker* | 3.78 (1.04-13.67) | **0.043** | 4.15 (1.1-15.68) | **0.036** |
| Stage* | 0.17 (0.03-1.14) | 0.069 | 0.1 (0.01-0.91) | **0.04** |
| Remission of LGD |  |  |  |  |
| Age at first resection* | 0.97 (0.91-1.02) | 0.25 | 0.98 (0.93-1.03) | 0.44 |
| Alcohol abuse (>14 units/per week)* | 2.12 (0.34-13.21) | 0.42 | 1.2 (0.18-7.99) | 0.85 |
| BMI | 1.05 (0.96-1.15) | 0.27 |  |  |
| Differentiation* | 0.63 (0.26-1.53) | 0.31 | 0.66 (0.26-1.65) | 0.37 |
| Lesion size (mm) | 0.95 (0.88-1.01) | 0.11 |  |  |
| Number of piecemeal resections | 0.83 (0.69-0.99) | 0.04 |  |  |
| Number of resections with OAC | 0.84 (0.66-1.05) | 0.12 |  |  |
| Percentage of resections with OAC**† | 0.98 (0.81-1.17) | 0.79 | 1.02 (0.84-1.24) | 0.84 |
| Sex* | 0.55 (0.13-2.36) | 0.42 | 0.34 (0.07-1.73) | 0.2 |
| Ever smoker* | 2.93 (1.05-8.16) | 0.04 | 3.39 (1.17-9.85) | **0.025** |
| Stage* | 0.3 (0.05-1.92) | 0.21 | 0.24 (0.03-1.84) | 0.17 |
| Remission of IM |  |  |  |  |
| Age at first resection* | 0.97 (0.93-1.01) | 0.16 | 0.98 (0.93-1.02) | 0.28 |
| Alcohol abuse (>14 units/per week)* | 1.34 (0.41-4.36) | 0.63 | 0.96 (0.28-3.37) | 0.95 |
| BMI | 1.03 (0.96-1.11) | 0.42 |  |  |
| Differentiation* | 0.69 (0.34-1.37) | 0.29 | 0.71 (0.34-1.48) | 0.36 |
| Lesion size (mm) | 0.99 (0.93-1.04) | 0.61 |  |  |
| Number of piecemeal resections | 0.97 (0.83-1.12) | 0.66 |  |  |
| Number of resections with OAC | 0.97 (0.79-1.19) | 0.78 |  |  |
| Percentage of resections with OAC**† | 0.98 (0.85-1.14) | 0.8 | 1.02 (0.87-1.19) | 0.82 |
| Sex* | 0.86 (0.32-2.35) | 0.77 | 0.75 (0.26-2.17) | 0.6 |
| Ever smoker* | 1.43 (0.67-3.05) | 0.36 | 1.52 (0.68-3.4) | 0.3 |
| Stage* | 0.26 (0.03-2.04) | 0.2 | 0.3 (0.04-2.49) | 0.26 |

**Supplementary Figure 1:**
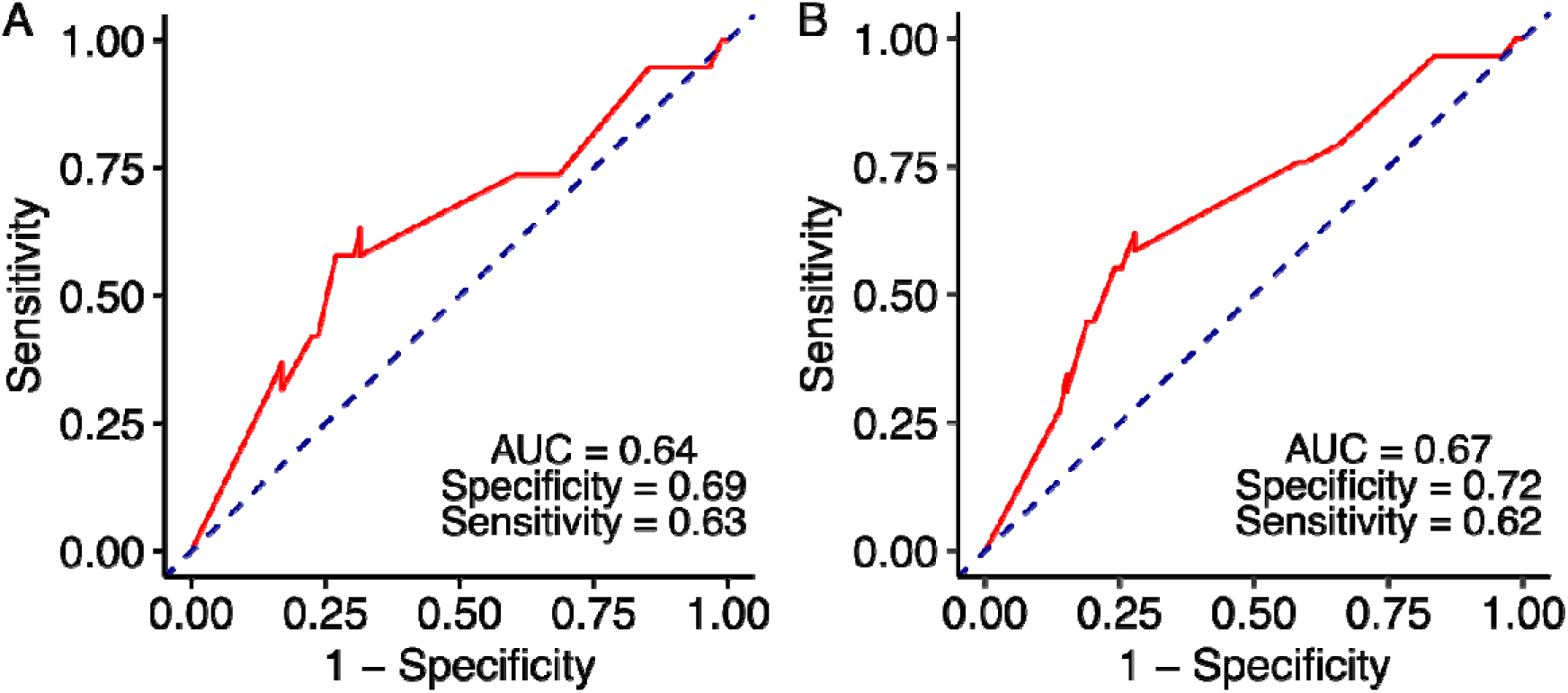
Receiver operating characteristics (ROC) curves. (A) ROC curve for prediction of risk of residual OAC based on the percentage of pEMR specimens with OAC at initial resection. Sensitivity and specificity are indicated for a 53.5% cut-off. (B) ROC curve for prediction of risk of recurrent OAC based on the percentage of pEMR specimens with OAC at initial resection. Sensitivity and specificity are indicated for a 53.5% cut-off.

## References

1. Morgan E, Soerjomataram I, Rumgay H, Coleman HG, Thrift AP, Vignat J, u. a. The Global Landscape of Esophageal Squamous Cell Carcinoma and Esophageal Adenocarcinoma Incidence and Mortality in 2020 and Projections to 2040: New Estimates From GLOBOCAN 2020. Gastroenterology [Internet]. 1. September 2022 [zitiert 25. Oktober 2024];163(3):649–658.e2. Verfügbar unter: http://www.gastrojournal.org/article/S0016508522006084/fulltext

2. Coleman HG, Xie SH, Lagergren J. The Epidemiology of Esophageal Adenocarcinoma. Gastroenterology. 1. Januar 2018;154(2):390–405.

3. Bhat S, Coleman HG, Yousef F, Johnston BT, McManus DT, Gavin AT, u. a. Risk of malignant progression in Barrett’s Esophagus patients: Results from a large population-based study. J Natl Cancer Inst. 6. Juli 2011;103(13):1049–57.

4. Shaheen NJ, Falk GW, Iyer PG, Souza RF, Yadlapati RH, Sauer BG, u. a. Diagnosis and Management of Barrett’s Esophagus: An Updated ACG Guideline. American Journal of Gastroenterology [Internet]. 1. April 2022 [zitiert 25. Oktober 2024];117(4):559–87. Verfügbar unter: https://journals.lww.com/ajg/fulltext/2022/04000/diagnosis_and_management_of_barrett_s_esophagus_.17.aspx

5. Weusten BLAM, Bisschops R, Dinis-Ribeiro M, di Pietro M, Pech O, Spaander MCW, u. a. Diagnosis and management of Barrett esophagus: European Society of Gastrointestinal Endoscopy (ESGE) Guideline. Endoscopy [Internet]. 28. November 2023 [zitiert 25. Oktober 2024];55(12):1124–46. Verfügbar unter: https://www.esge.com/diagnosis-and-management-of-barrett-esophagus

6. Di Pietro M, Trudgill NJ, Vasileiou M, Longcroft-Wheaton G, Phillips AW, Gossage J, u. a. National Institute for Health and Care Excellence (NICE) guidance on monitoring and management of Barrett’s oesophagus and stage I oesophageal adenocarcinoma. Gut [Internet]. 1. Juni 2024 [zitiert 25. Oktober 2024];73(6):897. Verfügbar unter: https://pmc.ncbi.nlm.nih.gov/articles/PMC11103346/

7. Wu J, Pan YM, Wang TT, Gao DJ, Hu B. Endotherapy versus surgery for early neoplasia in Barrett’s esophagus: a meta-analysis. Gastrointest Endosc [Internet]. 2014 [zitiert 25. Oktober 2024];79(2). Verfügbar unter: https://pubmed.ncbi.nlm.nih.gov/24079410/

8. Phoa KN, Pouw RE, Bisschops R, Pech O, Ragunath K, Weusten BLAM, u. a. Multimodality endoscopic eradication for neoplastic Barrett oesophagus: results of an European multicentre study (EURO-II). Gut [Internet]. 1. April 2016 [zitiert 25. Oktober 2024];65(4):555–62. Verfügbar unter: https://pubmed.ncbi.nlm.nih.gov/25731874/

9. Weusten BLAM, Bisschops R, Dinis-Ribeiro M, di Pietro M, Pech O, Spaander MCW, u. a. Diagnosis and management of Barrett esophagus: European Society of Gastrointestinal Endoscopy (ESGE) Guideline. Endoscopy [Internet]. 28. November 2023 [zitiert 9. Mai 2024];55(12):1124–46. Verfügbar unter: https://pubmed.ncbi.nlm.nih.gov/37813356/

10. Younis F, Rösch T, Beyna T, Ebigbo A, Faiss S, May A, u. a. Expert assessment of infiltration depth and recommendation of endoscopic resection technique in early Barrett cancer. United European Gastroenterol J [Internet]. 1. September 2024 [zitiert 1. Juli 2025];12(7). Verfügbar unter: https://pubmed.ncbi.nlm.nih.gov/38873843/

11. Gupta S, Mandarino FV, Shahidi N, Hourigan LF, Messmann H, Wallace MB, u. a. Can optical evaluation distinguish between t1a and t1b esophageal adenocarcinoma: an international expert inter-observer agreement study. Endoscopy [Internet]. 21. März 2024 [zitiert 1. Juli 2025];57(3). Verfügbar unter: https://pubmed.ncbi.nlm.nih.gov/39168143/

12. Terheggen G, Horn EM, Vieth M, Gabbert H, Enderle M, Neugebauer A, u. a. A randomised trial of endoscopic submucosal dissection versus endoscopic mucosal resection for early Barrett’s neoplasia. Gut [Internet]. 22. Januar 2017 [zitiert 25. Oktober 2024];66(5):783–93. Verfügbar unter: https://pubmed.ncbi.nlm.nih.gov/26801885/

13. Seewald S, Ang TL, Pouw RE, Bannwart F, Bergman JJ. Management of Early-Stage Adenocarcinoma of the Esophagus: Endoscopic Mucosal Resection and Endoscopic Submucosal Dissection. Dig Dis Sci [Internet]. 1. August 2018 [zitiert 25. Oktober 2024];63(8):2146–54. Verfügbar unter: https://link.springer.com/article/10.1007/s10620-018-5158-5

14. Guo HM, Zhang XQ, Chen M, Huang SL, Zou XP. Endoscopic submucosal dissection vs endoscopic mucosal resection for superficial esophageal cancer. World Journal of GastroenterologyL: WJG [Internet]. 14. Mai 2014 [zitiert 25. Oktober 2024];20(18):5540. Verfügbar unter: https://pmc.ncbi.nlm.nih.gov/articles/PMC4017070/

15. Martelli MG, Duckworth L V., Draganov P V. Endoscopic submucosal dissection is superior to endoscopic mucosal resection for histologic evaluation of barrett’s esophagus and barrett’s-related neoplasia. American Journal of Gastroenterology [Internet]. 1. Juni 2016 [zitiert 6. November 2024];111(6):902–3. Verfügbar unter: https://journals.lww.com/ajg/fulltext/2016/06000/endoscopic_submucosal_dissection_is_superior_to.41.aspx

16. Gray NA, Odze RD, Spechler SJ. Buried Metaplasia After Endoscopic Ablation of Barrett’s Esophagus: A Systematic Review. Am J Gastroenterol [Internet]. November 2011 [zitiert 9. November 2024];106(11):1899. Verfügbar unter: https://pmc.ncbi.nlm.nih.gov/articles/PMC3254259/

17. Titi M, Overhiser A, Ulusarac O, Falk GW, Chak A, Wang K, u. a. Development of Subsquamous High-Grade Dysplasia and Adenocarcinoma After Successful Radiofrequency Ablation of Barrett’s Esophagus. Gastroenterology [Internet]. 2012 [zitiert 9. November 2024];143(3):564. Verfügbar unter: https://pmc.ncbi.nlm.nih.gov/articles/PMC5511030/

18. Castela J, Serrano M, Ferro SM De, Pereira DV, Chaves P, Pereira AD. Buried Barrett’s Esophagus with High-Grade Dysplasia after Radiofrequency Ablation. Clin Endosc [Internet]. 1. Mai 2018 [zitiert 9. November 2024];52(3):269. Verfügbar unter: https://pmc.ncbi.nlm.nih.gov/articles/PMC6547340/

19. van Munster SN, Nieuwenhuis E, Bisschops R, Willekens H, Weusten BLAM, Herrero LA, u. a. Dysplastic Recurrence After Successful Treatment for Early Barrett’s Neoplasia: Development and Validation of a Prediction Model. Gastroenterology [Internet]. 1. Juli 2022 [zitiert 5. November 2024];163(1):285–94. Verfügbar unter: http://www.gastrojournal.org/article/S0016508522002712/fulltext

20. Pech O, Behrens A, May A, Nachbar L, Gossner L, Rabenstein T, u. a. Long-term results and risk factor analysis for recurrence after curative endoscopic therapy in 349 patients with high-grade intraepithelial neoplasia and mucosal adenocarcinoma in Barrett’s oesophagus. Gut [Internet]. 1. September 2008 [zitiert 21. Februar 2025];57(9):1200–6. Verfügbar unter: https://gut.bmj.com/content/57/9/1200

21. Gallegos MMM, Gomes ILC, Brunaldi VO, Bestetti AM, Marques SB, Miyajima NT, u. a. Endoscopic submucosal dissection vs. endoscopic mucosal resection in the treatment of early Barrett’s neoplasia: Systematic review and meta-analysis. Digestive Endoscopy [Internet]. 2024 [zitiert 6. November 2024]; Verfügbar unter: https://onlinelibrary.wiley.com/doi/full/10.1111/den.14892

22. Mejia Perez LK, Yang D, Draganov P V., Jawaid S, Chak A, Dumot J, u. a. Endoscopic submucosal dissection vsendoscopic mucosal resection for early Barrett’s neoplasia in the West: A retrospective study. Endoscopy [Internet]. 1. Mai 2022 [zitiert 5. November 2024];54(5):439–46. Verfügbar unter: http://www.thieme-connect.com/products/ejournals/html/10.1055/a-1541-7659

23. Komeda Y, Bruno M, Koch A. EMR is not inferior to ESD for early Barrett’s and EGJ neoplasia: An extensive review on outcome, recurrence and complication rates. Endosc Int Open [Internet]. 7. Mai 2014 [zitiert 6. November 2024];2(02):E58–64. Verfügbar unter: http://www.thieme-connect.com/products/ejournals/html/10.1055/s-0034-1365528

24. Doumbe-Mandengue P, Pellat A, Belle A, Ali EA, Hallit R, Beuvon F, u. a. Endoscopic submucosal dissection versus endoscopic mucosal resection for early esophageal adenocarcinoma. Clin Res Hepatol Gastroenterol. 1. Juni 2023;47(6):102138.

